# Acute kidney injury after liver resection: A systematic review, meta-analysis and metaregression of factors affecting it

**DOI:** 10.1101/2021.08.09.21261795

**Authors:** Bhavin Vasavada, Hardik Patel

**Affiliations:** Consultant hepatobiliary and liver transplant surgeon, Department of hepatobiliary and liver transplant surgery, Shalby Hospitals, Ahmedabad

**Keywords:** Acute kidney injury, liver resection, hepatectomy, meta-analysis, metaregression

## Abstract

**Aim:** This systematic review and meta-analysis aimed to study the incidence of acute kidney injury after liver resection and to analyze various factors affecting it by metaregression analysis.

**Methods:** The study was conducted according to the Preferred Reporting Items for Systematic Reviews and Meta-Analyses (PRISMA) statement (2020) and MOOSE guidelines. The meta-analysis was done using Review Manager 5.4 and the JASP Team (2020). JASP (Version 0.14.1)(University of Amsterdam). Weighted percentage incidence with 95% confidence intervals were used. Univariate metaregression was done by DerSimonian-Laird methods. Factors with a p-value less than 0.05 in the univariate metaregression model were entered in the multivariate metaregression model. Heterogeneity was assessed using the Higgins I^2^ test. The random-effects model was used in meta-analysis.

**Results:** Total 14 studies including 15510 patients were included in the final analysis. 1247 patients developed Acute Kidney Injury. Weighted Acute kidney injury percentage after liver resection was 15% with a 95% confidence interval of 11%-19%. On univariate metaregression analysis major hepatectomy (p=0.001), Underlying cirrhosis of liver (p=0.031), AKIN definition used (0.017), male sex (p<0.001), open surgery (p=0.032), underlying diabetes (0.026). On multivariate metaregression analysis major hepatectomy (p=0.003), underlying cirrhosis (p<0.001), male sex (p<0.001), AKIN classification used for defining acute kidney injury (p < 0.001, independently predicted heterogeneity and hence acute kidney injury.

**Conclusion:** Liver resection is associated with a high incidence of acute kidney injury. Major hepatectomy, male sex, underlying cirrhosis were independently predicting acute kidney injury.

## Introduction

Liver resections or partial hepatectomies are sometimes the only chance of cure in various benign and malignant conditions. Liver resections are still associated with high postoperative complications and mortality. [1,2,3]. The post-operative acute kidney is very common after liver resections. Different literature mentioned different incidences of acute kidney injury in various cohorts.[4].

Acute kidney injury incidence in overall noncardiac surgeries is around 1%, but it is very high after liver resections. However, the different article describes the different incidence of acute kidney injury after liver resection with incidence from 0.9% to around 20%. [5]. Incidence of postoperative acute kidney injury also depends upon various patient, tumor, and liver-related factors.

One of the reasons for heterogeneity in the literature is the variation in the definition of acute kidney injury used in the literature. Some used Acute Kidney Injury Network classification (AKIN), some used Risk, Injury, Failure, Loss and End-stage renal disease (RIFLE criteria) or some studies used Kidney Disease Improving Global Outcomes criteria (KDIGO) which combines RIFLE and AKIN criteria. [6,7,8].

### Aim

This systematic review and meta-analysis aimed to study the prevalence of acute kidney injury after liver resection and to analyze various factors affecting it by metaregression analysis.

## Methods

The study was conducted according to the Preferred Reporting Items for Systematic Reviews and Meta-Analyses (PRISMA) statement (2020) and MOOSE guidelines. [9,10]. We conducted a literature search as described by Gossen et al. PubMed, Cochrane Library, Embase, google scholar, web of science with mesh terms “Acute kidney injury” AND “liver resection” OR “hepatectomy”. [11] Two independent authors extracted the data. In case of disagreements, decisions are reached on basis of discussions. We defined major hepatectomy as >= 3 liver segments. We defined chronic renal failure as preoperative eGFR < 90 ml/min/1.73m2.

### Statistical Analysis

The meta-analysis was done using Review Manager 5.4 and the JASP Team (2020). JASP (Version 0.14.1)(University of Amsterdam). Weighted percentage incidence with 95% confidence intervals were used. Univariate metaregression was done by DerSimonian-Laird methods. Factors with a p-value less than 0.05 in the univariate metaregression model were entered in the multivariate metaregression model. Heterogeneity was assessed using the Higgins I^2^ test [12], with values of 25%, 50%, and 75% indicating low, moderate, and high degrees of heterogeneity, respectively, and assessed p-value for the significance of heterogeneity and tau^2^ and H^2^ value. The random-effects model was used in meta-analysis.

### Assessment of Bias

Cohort studies were assessed for bias using the Newcastle-Ottawa Scale to assess for the risk of bias Publication bias was assessed using a funnel plot. [13,14]. Funnel plot asymmetry was evaluated by Egger’s test.

### Inclusion and Exclusion criteria for studies

Inclusion criteria:

- Studies with full texts
- Studies that evaluated acute kidney injury after liver resection
- English language studies

Exclusion criteria:

- Studies which were not fulfilling the above criteria.
- Duplicate studies.

## Results

### Data extraction, study characteristics, and quality assessment

‘PUBMED’, ‘SCOPUS’, and ‘EMBASE’ databases were searched using keywords and the search strategy described above. 31700 studies were found using earlier mentioned MESH terms after duplicates were removed 30573 studies were screened, 30556 studies were excluded after applying exclusion criteria. Out of 17 studies remaining full texts of 3 studies could not be retrieved and excluded. 14 studies including 15510 patients who underwent liver resections were included in the final analysis. [Figure 1]. The risk of bias summary is mentioned in Figure 2. Study characteristics are mentioned in table 1. All studies were either retrospective cohort, prospective cohort, or propensity score-matched study. In propensity score-matched analysis unmatched total data were analyzed for postoperative morbidity.

**Figure 1:**
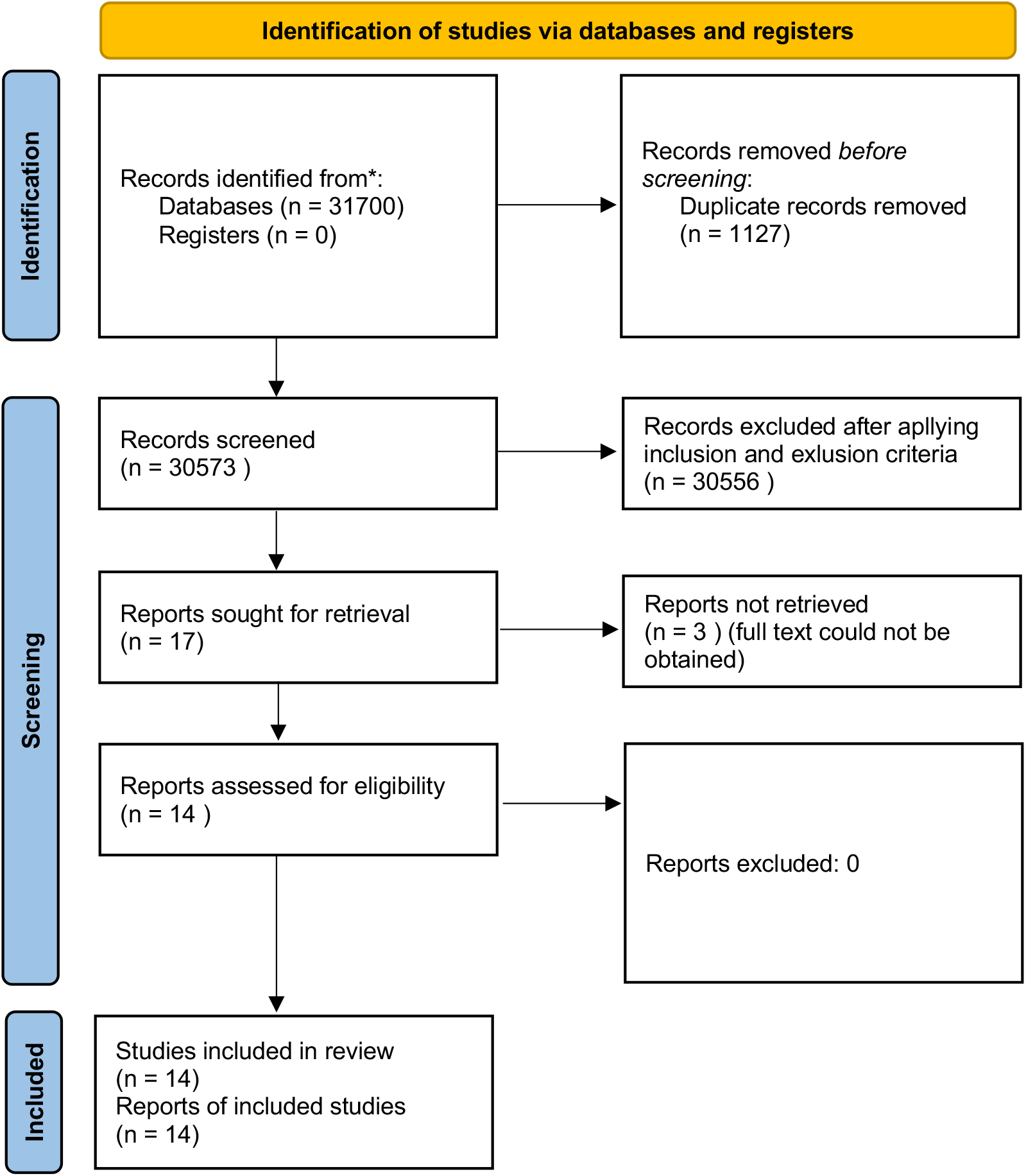
Prisma flow chart **PRISMA 2020 flow diagram for new systematic reviews which included searches of databases and registers only**

**Figure 2:**
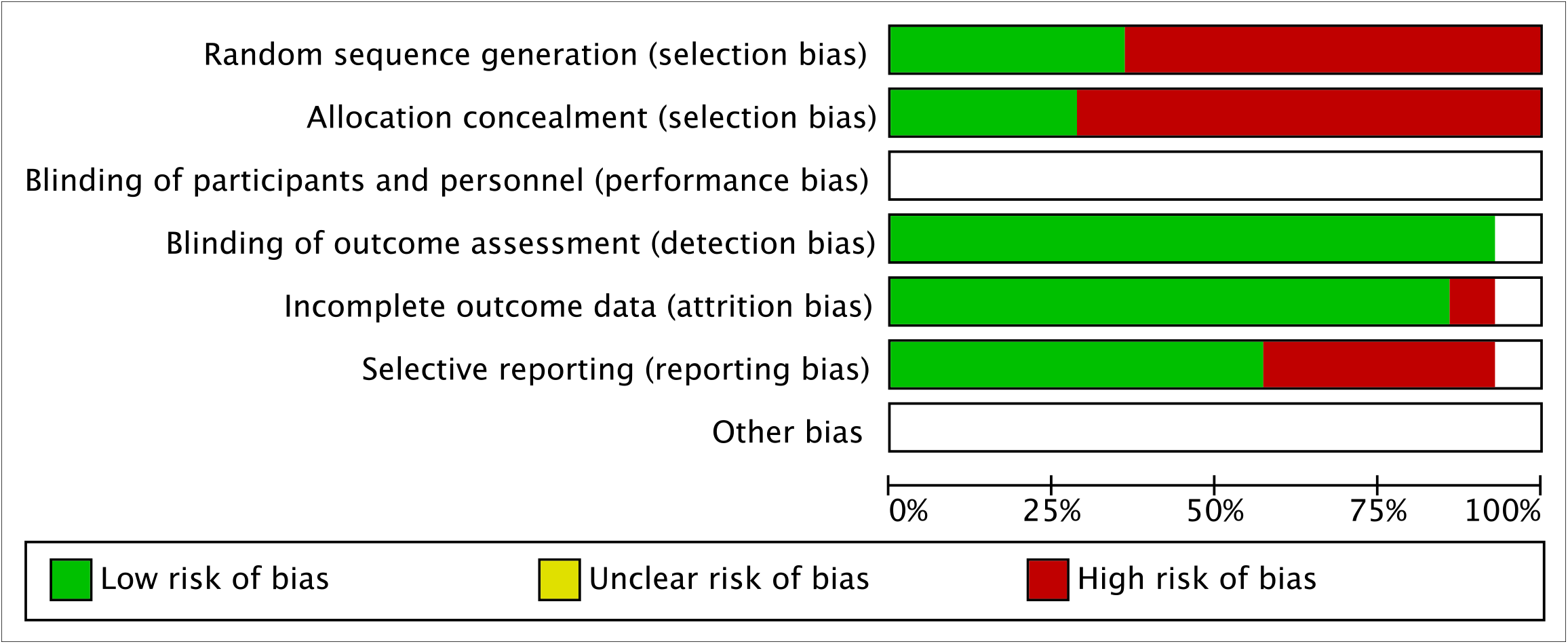
Risk of bias summary

### Acute Kidney Injury

Total 14 studies including 15510 patients were included in the final analysis. [15-28]. 1247 patients developed Acute Kidney Injury. Weighted Acute kidney injury percentage after liver resection was 15% with a 95% confidence interval of 11%-19% as shown in the forest plot. [Figure 3]. 5 studies used AKIN criteria, 6 used KDIGO, and 1 used RFILE criteria, for two studies detailed of Acute kidney injury definitions were not available. [Table 1]. However, the heterogeneity of the analysis was high with I2 98.83 % (p<0.01). Publication bias was also significant with a p-value <0.01, as shown in the funnel plot. [figure 4]

**Figure 3:**
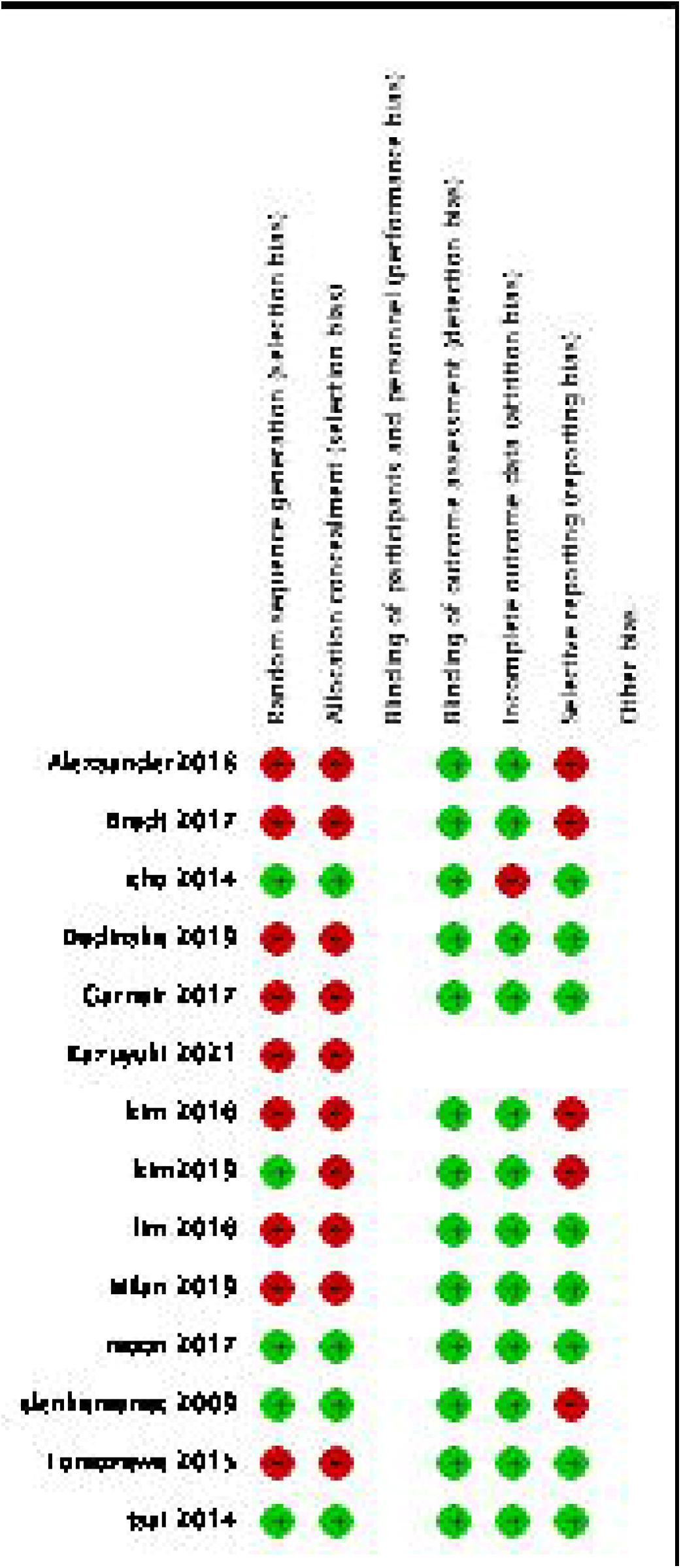
Forest plot for acute kidney injury

**Figure 4:**
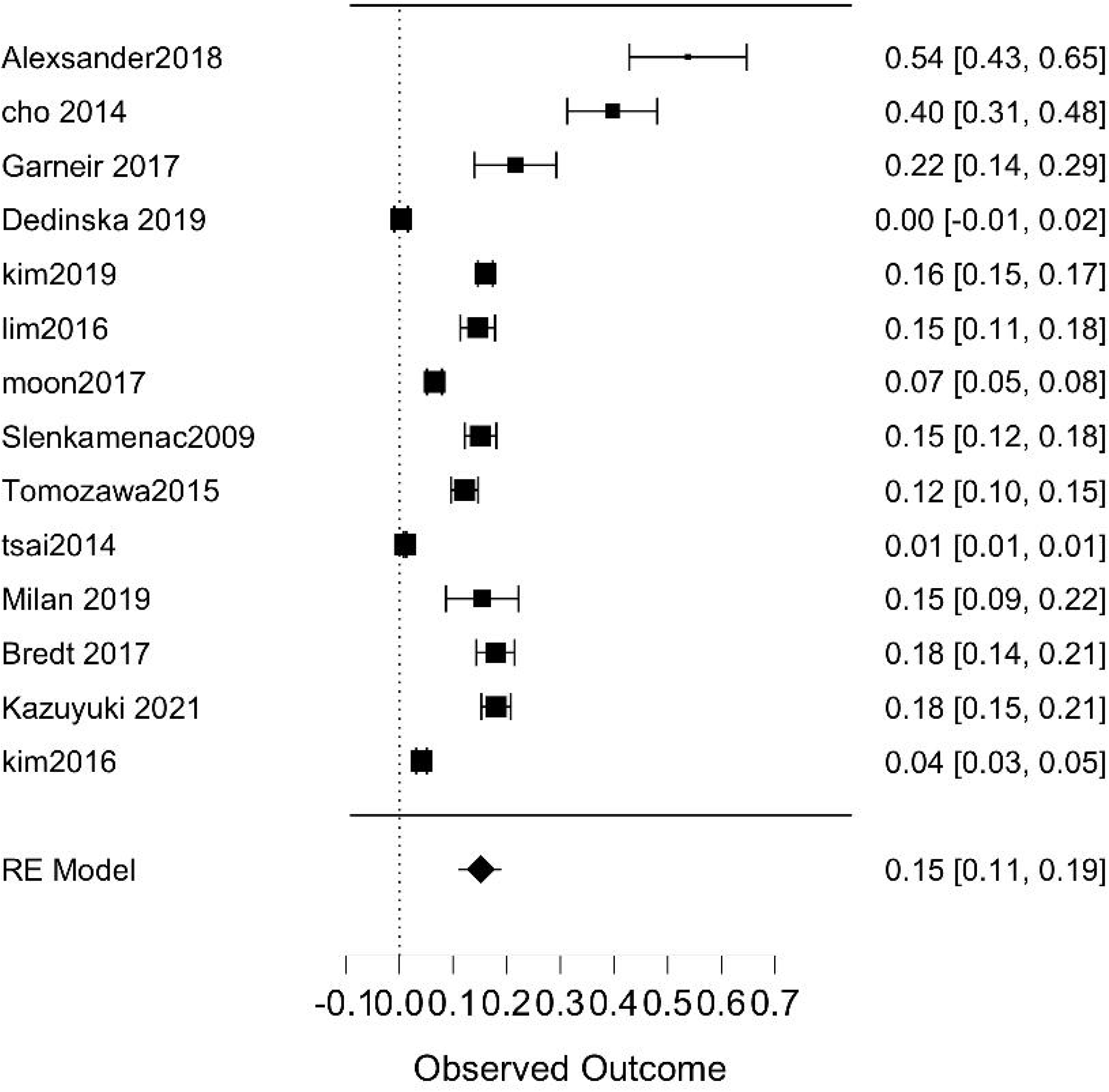
Funnel plot for publication bias

Meta-regression analysis: [Table 2].

On univariate metaregression analysis major hepatectomy (p=0.001), Underlying cirrhosis of liver (p=0.031), AKIN definition used (0.017), male sex (p<0.001), open surgery (p=0.032), underlying diabetes (0.026). On multivariate metaregression analysis major hepatectomy (p=0.003), underlying cirrhosis (p<0.001), male sex (p<0.001), AKIN classification used for defining acute kidney injury (p < 0.001, independently predicted heterogeneity and hence acute kidney injury. Residual heterogeneity after multivariate metaregression analysis was nonsignificant (p=0.065) and publication bias with eagers’ test was also nonsignificant. (p= 0.292). Multivariate metaregression forest plot and funnel plot for publication bias is shown in supplement figure 1.

## Discussion

Acute kidney injury post liver resection is a serious problem and is always associated with higher postoperative mortality. [1,2,3]. Many individual cohort studies are available to know the incidence of acute kidney injury after liver resection. However, there was a wide difference in incidence rates, one of the reasons for it may be variation in techniques, patient population, type of surgeries, and because of that, we decided to perform systematic review and metaanalysis with metaregression analysis to look for reasons of heterogeneity and factors responsible for heterogeneity and in the process with acute kidney injury, and also to find the pooled prevalence of acute kidney injury post-liver resections.

The weighted pooled prevalence of acute kidney injury after liver resection was 15% (95% confidence interval 11-19%), however as expected heterogeneity was significant and high with I2 of 98.83%, suggesting a difference in Acute kidney injury according to the various study population. After multivariate metaregression analysis major hepatectomy, underlying cirrhosis, male sex and AKIN classification used were independently associated with heterogeneity and hence acute kidney injury and difference acute kidney injury rates across various centers.

The fact that Acute Kidney Injury Network or AKIN definition of acute kidney injury independently predicted heterogeneity across various studies shows that acute kidney injury incidence was different across the studies differed somewhat due to definitions used and it shows the need for a standard definition of postoperative acute kidney injury as some decrease in urine output is expected after major abdominal surgery due to increased fluid shifts and third space loss in post-operative periods.

Amount of intraoperative fluids, intraoperative hypotension, blood loss were not associated with heterogeneity in the acute kidney injury incidences in our metanalysis which may suggest the perioperative practices are standardized across the centers. Preoperative diabetes was associated with heterogeneity in univariate analysis but failed to predict heterogeneity independently in multivariate metaregression analysis. Preoperative hypertension and pre-existing chronic renal failure were not associated with heterogeneity across the centers.

There are some limitations in our analysis, as most of the studies were retrospective, so, we could not entirely rule out the selection and reporting bias as shown in the summary of bias. The strength of this analysis was that to our knowledge this is the first metanalysis with metaregression analysis studying various factors associated with heterogeneity across the studies and hence acute kidney injury.

## Conclusion

Liver resection is associated with a high incidence of acute kidney injury. Major hepatectomy, male sex, underlying cirrhosis were independently predicting acute kidney injury. Acute kidney injury incidence also varied according to the acute kidney injury definition used.

## Data Availability

data will be made available on demand

## Abbreviations

(AKIN): Acute Kidney Injury Network classification
(RIFLE): Risk, Injury, Failure, Loss and End stage renal disease
(KDIGO): Kidney Disease Improving Global Outcomes criteria

## Figure legends

**Supplement figure 1:**
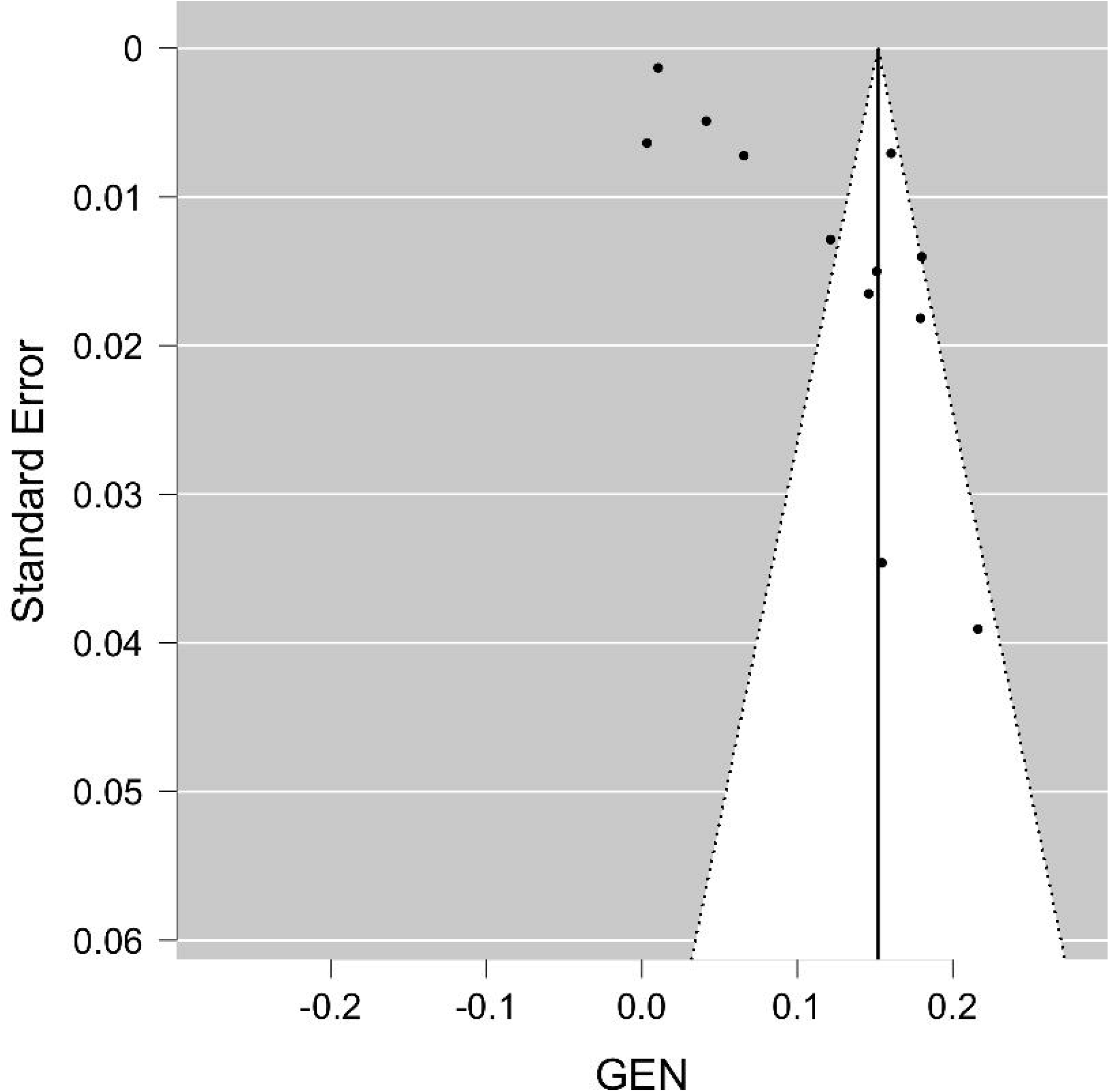
Metaregression forest plot and funnel plot.

