## Supplementary material for "Acute kidney injury after liver resection: A systematic review, meta-analysis and metaregression of factors affecting it": table 1

| **study** | **Type of Study** | **AKI (n)** | **AKI (%)** | **Total Patients(n)** | **Major Hepatectomy** | **intraoperative hypotension** | **Cirrhosis** | **Operative time (minutes)** | **Blood loss (ml)** | **Akin Classification** | **Sex (male) (n)** | **Open Surgery(n)** | **Diabetes** | **Age (mean)** | **Intra operative fluid (ml)** | **Hypertension** | **Kidigo Criteria** | **Chronic Renal Failure** | **RIFILE Criteria** |
| --- | --- | --- | --- | --- | --- | --- | --- | --- | --- | --- | --- | --- | --- | --- | --- | --- | --- | --- | --- |
| **Alexsander2018 [15]** | **Retrospective cohort** | 43 | 53 | 80 | 20 |  | 64 |  | 250 | Yes | 61 | 67 | 20 | 62 |  | 28 | No | 44 | No |
| **cho 2014 [16]** | **Retrospective cohort** | 52 | 40 | 131 | 100 | 34 | 7 | 300 | 350 | Yes | 82 | 131 | 32 | 56 | 1985 | 44 | No | 0 | No |
| **Garneir 2017 [17]** | **Retrospective cohort** | 24 | 21.6 | 111 | 106 | 46 | 2 | 340 | 500 | No | 55 | 109 | 13 | 66 | 3000 |  | No | 11 | No |
| **Dedinska 2019 [18]** | **Retrospective cohort** | 26 | 3 | 785 | 107 |  |  |  |  | No |  |  |  | 58.7 |  |  | Yes |  | No |
| **kim2019 [19]** | **Retrospective cohort** | 432 | 16.04 | 2692 | 918 |  | 257 | 250 |  | No | 1358 | 2692 | 444 | 60 |  | 1193 | Yes | 1154 | No |
| **lim2016 [20]** | **Retrospective cohort** | 67 | 14.6 | 457 | 241 |  | 206 | 208 |  | No | 379 | 329 | 75 | 60.5 |  |  | Yes | 70 | No |
| **moon2017 [21]** | **Propensity score matched** | 77 | 6.56 | 1173 | 790 |  |  | 271 |  | No | 951 | 926 | 75 | 55.7 | 2263 | 80 | Yes | 866 | No |
| **Slenkamenac2009 [22]** | **Prospective cohort** | 86 | 15.11 | 569 | 326 |  | 40 | 294.2 | 551 | No | 311 | 573 | 59 | 57.2 |  |  | No | 73 | Yes |
| **Tomozawa2015 [23]** | **Retrospective cohort** | 78 | 12.14 | 642 | 230 |  | 93 | 300 | 1310 | Yes | 473 | 642 | 130 | 67 | 4400 | 310 | No | 124 | No |
| **tsai2014 [24]** | **Retrospective cohort** | 62 | 10.46 | 5924 | 1140 |  | 1957 |  |  | No | 4273 |  | 2962 | 63 |  | 577 | No | 80 | No |
| **Milan 2019 [25]** | **Retrospective cohort** | 17 | 15.45 | 109 | 46 |  |  | 233 |  | No | 55 | 92 | 26 | 61 |  |  | Yes |  | No |
| **Bredt 2017 [26]** | **Retrospective cohort** | 80 | 17.93 | 446 | 128 | 26 | 23 |  |  | Yes | 223 | 343 | 44 | 54.6 |  | 70 | No |  | No |
| **Kazuyuki 2021 [27]** | **Retrospective cohort** | 135 | 18 | 750 | 285 |  | 111 | 330 | 300 | No |  | 540 | 194 |  | 2550 | 385 | Yes | 569 | No |
| **kim2016 [28]** | **Propensity score matched** | 68 | 4.14 | 1641 | 1641 |  | 0 |  |  | Yes | 1109 | 1641 | 0 | 27.5 |  | 0 | No | 0 | No |

Table 1. Patients’ characteristics.
