## Supplementary material for "Acute kidney injury after liver resection: A systematic review, meta-analysis and metaregression of factors affecting it": table 2

| Factors | Univariate metaregression analysis (p value) | Multivariate metaregression analysis (p value) |
| --- | --- | --- |
| Major hepatectomy | **0.001** | **0.003** |
| Intra operative hypotension | 0.06 |  |
| Underlying cirrhosis | **0.031** | **<0.001** |
| Operative time | 0.279 |  |
| Blood loss | 0.232 |  |
| AKIN classification | **0.017** | **<0.001** |
| Male sex | **<0.001** | **<0.001** |
| Open hepatectomy | **0.032** | 0.217 |
| Child pugh’s score | 0.298 |  |
| Diabetes | **0.026** | 0.180 |
| Age | 0.201 |  |
| Intra operative fluids (ml) | 0.364 |  |
| Hypertension | 0.234 |  |
| KDIGO criteria | 0.102 |  |
| Previous chronic renal failure | 0.292 |  |
| RIFLE criteria | 0.994 |  |

Table 2: Univariate and multivariate metaregression analysis. (All factors with p value less than 0.05 were entered in multivariate analysis)
