## supplement figure 1 for "Acute kidney injury after liver resection: A systematic review, meta-analysis and metaregression of factors affecting it"

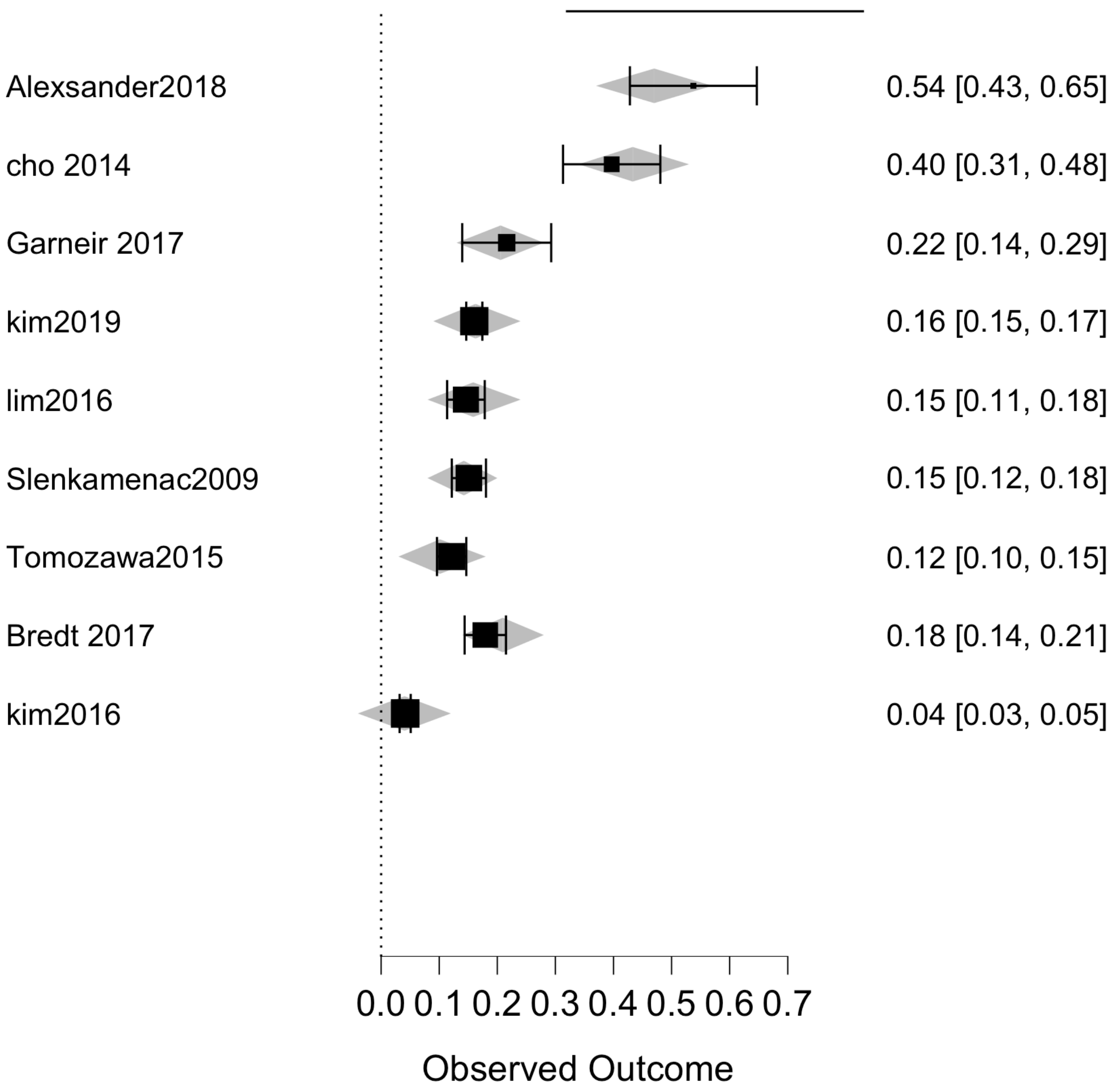


Supplement Figure 1 (a) Metaregression forest plot.

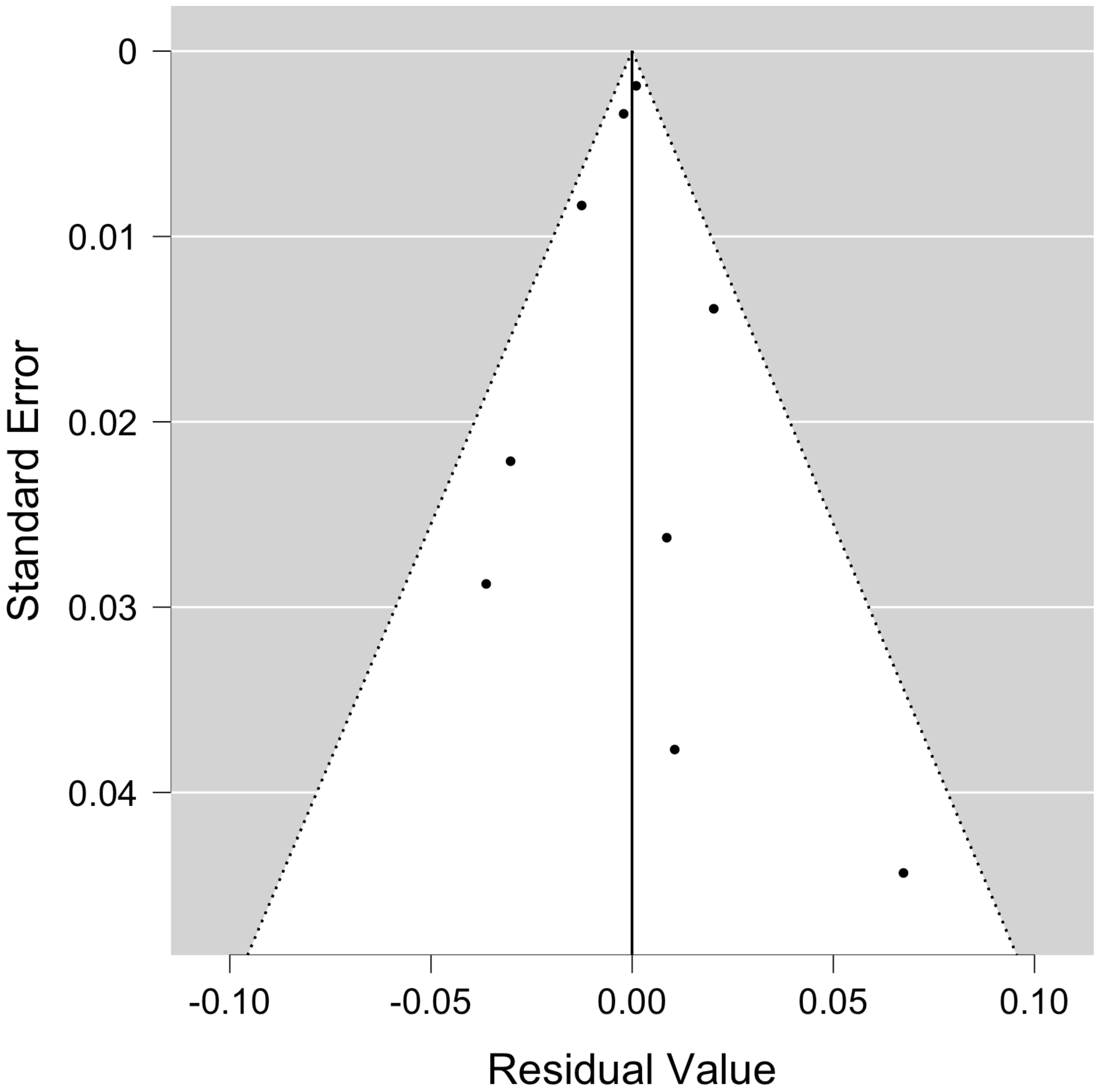


Supplement figure 1 (b) Metaregression funnel plots.
